# Longitudinal dynamics of respiratory microbiome composition in infants after tracheostomy placement

**DOI:** 10.64898/2025.12.29.25342834

**Authors:** Rebecca Steuart, Samantha N. Atkinson, Lucas R. Hoffman, Long Hung, Xiangming Ding, Nita Salzman, Christopher J. Russell

## Abstract

Abnormal respiratory microbiomes are reported in children with artificial airways, yet the timing and persistence of these disruptions have not been defined in infants following new tracheostomy placement. We conducted a prospective longitudinal study to characterize airway microbiome dynamics following new tracheostomy placement during early life, a critical period for microbiome development. Fifteen hospitalized infants <=12 months contributed 84 tracheal aspirate samples collected from day 1 through 3 to 4 months post-procedure. 16S rRNA sequencing revealed immediate and sustained community shifts. *Staphylococcus* abundance increased after tracheostomy, peaking at 40 days (mean 27%) before declining, with a more pronounced bloom in infants without home mechanical ventilation (HMV). Alpha diversity decreased significantly in the first 30 days (p<0.05) and returned to baseline by 61 to 90 days. Beta diversity analysis demonstrated marked compositional changes immediately post-tracheostomy and ongoing divergence through 3 to 4 months. Time since tracheostomy and clinical factors (gestational age, HMV, neurologic impairment) were significantly associated with microbiome structure (p=0.001). These findings provide novel evidence that tracheostomy induces rapid and prolonged airway microbiome disruption in infants, highlighting a previously uncharacterized window of vulnerability with implications for respiratory health and individualized care.

## Introduction

There is emerging evidence for abnormal respiratory microbiomes among children with artificial airways, including both new and long-term tracheostomies and endotracheal tubes.^1,2^ However, the temporal dynamics of that dysbiosis, including onset and chronicity, have not been well-described. Defining these dynamics for infants with new tracheostomies, and their causes and consequences, could help identify strategies to promote microbiome health and clinical stability, especially during the critical period of microbiome development in early life.

In this study, we aimed to identify the temporal dynamics and clinical associations of tracheostomy aspirate (TA) microbiomes among a cohort of infants followed longitudinally after new tracheostomy placement.

## Methods

This was a prospective study at Children’s Hospital Los Angeles from 2020-2021. We enrolled hospitalized infants ≤12 months old undergoing new tracheostomy placement and collected serial TA samples on day 1, at first tracheostomy change (day 7), and every ∼2 weeks thereafter during initial hospitalization. Samples were collected immediately following tracheostomy changes and stored at -80°C. Clinical characteristics were extracted from medical records. Institutional review board approval was obtained and parents provided written informed consent.

DNA was extracted from samples using QIAamp PowerFecal Pro DNA Kits (Qiagen) via the Qiacube platform using manufacturer’s protocol. The V4 region of 16S bacterial DNA was PCR-amplified using shared forward primer 806rB; each sample had unique reverse primers.^3,4^ Samples were pooled and sequenced using custom primers by paired-end Illumina MiSeq(2x150bp). Reads were processed using DADA2v1.5.2 and taxonomies assigned using RDP naïve Bayes classifier. Diversity metrics were computed using genus taxa counts.

We used a linear mixed-effects model to identify differences in each infant’s change from baseline in Shannon alpha diversity over time, including fixed effects for use of chronic or home mechanical ventilation (HMV, defined as ventilator use at discharge), gestational age category, sex, age at tracheostomy, presence of neurologic impairment, upper airway obstruction, and craniofacial diagnoses, as well as a random intercept for infant. Principal coordinates analysis (PCoA) plots with a multivariable PERMANOVA model evaluated associations between time with tracheostomy and Bray-Curtis dissimilarity, accounting for key clinical factors from marginal testing and repeated measures with stratification. Analyses used ‘phyloseq’(v1.20.0) and ‘vegan’(v2.5-4) in R v4.4.1.

## Results

Among the 15 infants included, most were female (60%) and Latino/a/x/Hispanic (60%) or Black (33%). Eight infants (53%) were preterm, and median gestational age was 36 weeks (IQR 32-38.5). Indications for tracheostomy included upper airway obstruction (40%), neurologic impairment (40%), craniofacial anomalies (20%), and bronchopulmonary dysplasia (20%), with some having multiple indications; 40% had other indication. Median age at tracheostomy was 5 months (IQR 2.5-7). All infants were discharged home, 67% using HMV. Acute respiratory illnesses were rare (n=1).

Median hospital stay and sampling duration was 67 days (IQR 29-118.5); 84 samples were of sufficient quality for analysis, with median 6 samples per infant (range 3-11). The most abundant genera identified included *Staphylococcus* (mean relative abundance 21%), *Streptococcus* (12%), *Klebsiella* (10%), *Neisseria* (10%), *Pseudomonas* (7%), *Stenotrophomonas* (7%), and *Serratia* (7%). Only 3 infants had samples with >1% relative abundance of *Pseudomonas*, all with HMV. *Staphylococcus* relative abundance increased following tracheostomy, peaking at 40 days and mean relative abundance of 27%, followed by a subsequent decrease; this was more pronounced among infants without HMV (57% vs. 20% peak).

Longitudinal analysis of alpha (within-sample) diversity using Shannon index identified a significant decrease in median alpha diversity from baseline in the first 30 days (p=0.017 Day 7-13; p=0.040 Day 13-30). By 61-90 days, median alpha diversity returned to baseline (p=0.11 Day 31-60; p=0.55 Day 61-90).

PCoA plots of beta (between-sample) diversity using Bray-Curtis dissimilarity demonstrated that samples collected immediately following tracheostomy clustered together (Day 1-2, **Figure 1**), indicating relatively similar microbiome compositions, followed by divergence by Day 14-30 and some return of clustering by Day 61-90. Multivariable PERMANOVA showed time post-tracheostomy was significantly associated with community structure when accounting for clinical variables (p=0.001). Biplots indicated Day 14-30 samples had higher abundances in *Staphylococcus*. Gestational age, HMV, neurologic impairment, and upper airway obstruction were also significantly associated with microbiome composition (all p=0.001). Over time, microbiome communities rapidly shifted into communities dissimilar to baseline, and continued to have ongoing composition changes with each successive sampling timepoint post-tracheostomy without stabilization or return to baseline (**Figure 2**).

**Figure 1.**
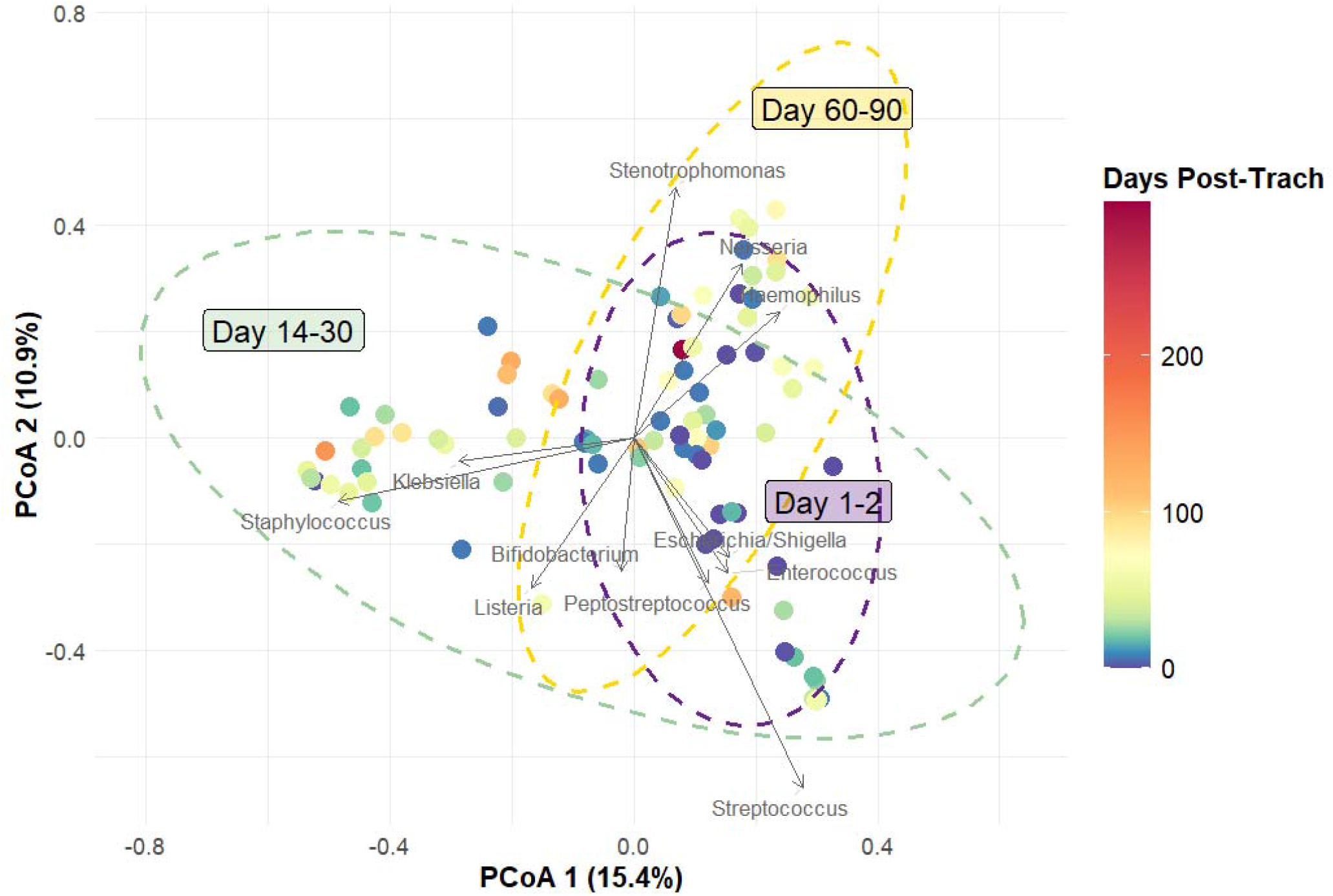
Tracheostomy aspirate microbiome communities diverge in the first month after tracheostomy placement.

**Figure 2.**
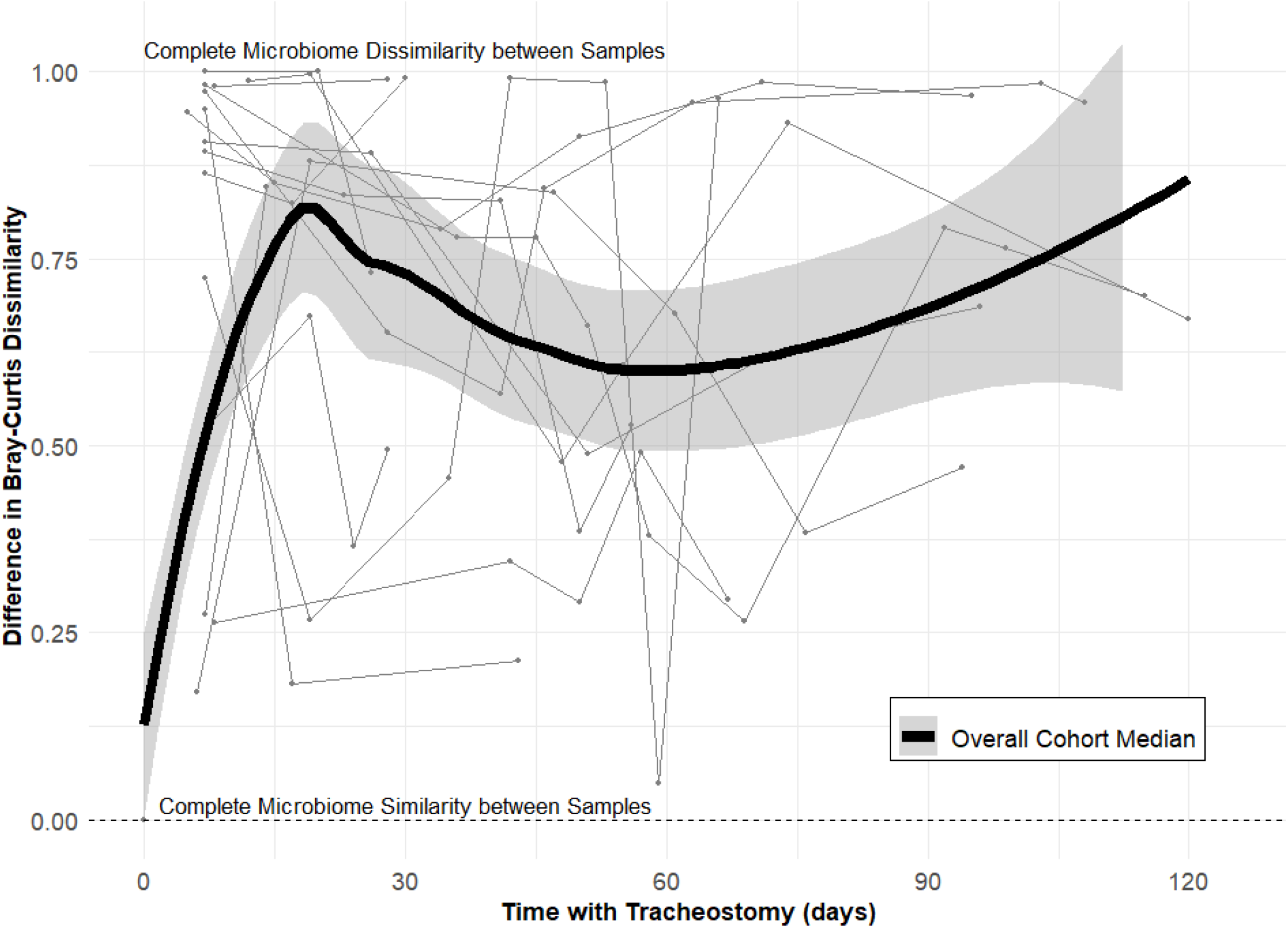
Tracheostomy aspirate microbiome communities have ongoing changes with each sampling timepoint during the first months after tracheostomy placement.

## Discussion

We identified significant shifts in respiratory sample microbiomes in infants within the first weeks after new tracheostomy, including a transient bloom in *Staphylococcus* abundance. Although bacterial richness and evenness largely returned to baseline, there were ongoing microbiome community composition changes through 3-4 months. These patterns occurred across gestational age and HMV status. Together, our findings suggest that tracheostomy placement leads to immediate and sustained microbiome community disruptions in the infant airway.

Among healthy infants, the oropharyngeal and lung microbiomes are dominated by *Streptococcus, Neisseria, Haemophilus*, and *Veillonella* after the first week of life.^5-7^ Therefore, the transient increase in *Staphylococcus* and declines in diversity that we observed likely reflect effects of tracheostomy tube placement, including entry of skin flora via the stoma, perioperative antibiotics, and altered mucociliary clearance. A similar increase in *Staphylococcus* was also reported in older children post-tracheostomy, and following intubation among infants with pneumonia.^1,2^ Blooms of other resident bacteria have also been described among children with tracheostomies during viral infection.^8^ Differential diversity recovery among gestational age and HMV groups suggests that these factors influence microbiological trajectories.

It is not known if the continual shifts in microbial communities we observed at 3 months persist. If so, this could have lasting implications for airway health, and as such could influence the occurrence and severity of respiratory illness given the respiratory microbiome’s role in shaping respiratory immunity and inflammatory responses.^1,9^ Understanding the role of these dynamic community structures in resilience or vulnerability will be critical for predicting disease trajectory and guiding individualized, long-term management strategies.

This study has several limitations, including a small patient cohort and single-center design, lack of control group, and potential residual confounding from unmeasured variables. These limitations are balanced by notable strengths: a longitudinal design, dense sampling, and low respiratory illness prevalence. Notably, our individual-level analysis contrasts with previous group-level studies, and underscores the importance of analyzing microbiome temporal dynamics at the individual level.^1,10^

In summary, tracheostomy placement leads to both short- and longer-term disruptions in the infant respiratory microbiome. Future work should explore the clinical outcomes associated with microbiome community alterations, potentially leading to targeted treatment approaches.

Principal coordinates analysis (PCoA) with Biplot vectors of tracheostomy aspirate microbial communities using Bray-Curtis dissimilarity index. Points represent individual sample microbiota; color represents days since tracheostomy. Ellipses indicate 95% confidence intervals for group centroids. Vectors represent bacterial genera that are significantly associated with microbial community composition (p<0.05).

Beta-diversity measures over time demonstrated clustering immediately following tracheostomy (Day 1-2, purple), followed by notable shifts by Day 14-30 (green) and again by Day 60-90 (yellow). Multivariable PERMANOVA analysis showed that time post-tracheostomy was significantly associated with bacterial community structure when accounting for other clinical variables (p=0.001). Biplots indicated that Day 1-2 communities had abundance of *Streptococcus* while Day 14-30 communities moved transiently to higher abundance of *Staphylococcus* and *Klebsiella*. Gestational age, chronic mechanical ventilation, neurologic impairment, and upper airway obstruction were also statistically significantly associated with bacterial composition (all p=0.001). Bronchopulmonary dysplasia diagnosis was not included in the model as it was collinear with neurologic impairment.

Successive sample differences in Bray-Curtis dissimilarity over time following tracheostomy. Lines represent differences in Bray-Curtis dissimilarity between successive samples for individual infants. Difference value of 0 indicates complete microbiome community similarity between an individual’s successive samples, while 1.0 indicates complete dissimilarity between samples. Gray shading denotes the 95% confidence interval of the Overall Cohort Median.

Within the first week post-tracheostomy, TA microbiome communities rapidly shifted into dissimilar compositions from baseline, as evidenced by Difference in Bray-Curtis values >0.50. Infants had ongoing community changes with each successive sampling timepoint, without achieving compositional similarity. Not pictured: After initial microbiome composition shifts post-tracheostomy, TA microbiome communities did not return to compositions similar to those of baseline either, but rather remained dissimilar throughout the sampling period.

## Data Availability

All data produced in the present study are available upon reasonable request to the authors.

## Abbreviations

TA: tracheostomy aspirate
IQR: interquartile range
HMV: home mechanical ventilation
PCoA: principal coordinates analysis
PERMANOVA: permutational analysis of variance

